# Excess mortality in England and Wales during the first wave of the COVID-19 pandemic

**DOI:** 10.1101/2020.05.26.20113357

**Authors:** Evangelos Kontopantelis, Mamas A Mamas, John Deanfield, Miqdad Asaria, Tim Doran

## Abstract

**Background:** Deaths during the COVID-19 pandemic result directly from infection and exacerbation of other diseases and indirectly from deferment of care for other conditions, and are socially and geographically patterned. We quantified excess mortality in regions of England and Wales during the pandemic, for all causes and for non-COVID-19 associated deaths.

**Methods:** Weekly mortality data for 1 Jan 2010 to 1 May 2020 for England and Wales were obtained from the Office of National Statistics. Mean-dispersion negative binomial regressions were used to model death counts based on pre-pandemic trends and exponentiated linear predictions were subtracted from: i) all-cause deaths; and ii) all-cause deaths minus COVID-19 related deaths for the pandemic period (07-13 March to 25 April to 8 May).

**Findings:** Between 7 March and 8 May 2020, there were 47,243 (95%CI: 46,671 to 47,815) excess deaths in England and Wales, of which 9,948 (95%CI: 9,376 to 10,520) were not associated with COVID-19. Overall excess mortality rates varied from 49 per 100,000 (95%CI: 49 to 50) in the South West to 102 per 100,000 (95%CI: 102 to 103) in London. Non-COVID-19 associated excess mortality rates ranged from −1 per 100,000 (95%CI: −1 to 0) in Wales (i.e. mortality rates were no higher than expected) to 26 per 100,000 (95%CI: 25 to 26) in the West Midlands.

**Interpretation:** The COVID-19 pandemic has had markedly different impacts on the regions of England and Wales, both for deaths directly attributable to COVID-19 infection and for deaths resulting from the national public health response.

**Funding:** None

**Summary box:** *What is already known on the subject:*

1. The number of deaths due to COVID-19 have been quantified by the Office of National Statistics
2. These have also been reported across age groups and regions

*What this study adds:*

1. We report the number of excess deaths, using weekly mortality data from 1/1/2010
2. We also quantify the number of excess deaths, excluding COVID-19 associated deaths, which can be attributed to COVID-19 directly (but not coded as such) or indirectly (due to other urgent but unmet health need)
3. Highest excess mortality, excluding COVID-19 deaths, was observed in the West Midlands, followed by London and the North West
4. Although males had larger excess mortality rates than females across all age groups, female excess mortality rates excluding COVID-19 were higher in the 85+ age group, indicating a large undocumented impact of the virus on older females (direct and/or indirect)
5. The three provided appendices will be updated weekly on the BMJ-JECH website, to provide up-to-date information of excess mortality by region, sex and age group

## Introduction

The COVID-19 pandemic began in Hubei Province, China towards the end of 2019, and the first confirmed cases in Britain were recorded on January 31 2020. As of 21 May 2020 there were 158,488 confirmed cases in England and Wales and 33,081 deaths in people testing positive for SARS-CoV-2.^1^ In addition to increased risks for men, older age groups,^2^ and people with long term conditions such as cardiovascular disease and diabetes, it was clear from the earliest stages of the pandemic that the consequences of the outbreak would be more severe for some social groups, generally those already most affected by health inequalities. In mid-April, deaths involving COVID-19 in England ranged from 25 per 100,000 in the most affluent 10% of areas to 55 per 100,000 in the most deprived 10%, following the same social gradient as pre-pandemic all cause death rates.^3^ Similar patterns emerged in Wales. Ethnic minority groups were also disproportionately affected, with mortality rates in for Bangladeshi and Pakistani ethnicity over three times higher than for White, and rates for Black ethnicity over four times higher.^4^ To some extent, these variations may reflect baseline health inequalities across society, but social deprivation also appears to increase susceptibility to COVID infection specifically, through factors including overcrowding, working in a key service or agricultural industry, or having serious underlying medical conditions that increase the risk of COVID-related mortality.

The COVID-19 pandemic has also generated geographic patterns of impact that interact in complex ways with social patterns. London, a major international gateway, was the first area to be substantially affected and as of mid-May was the region with the highest number of confirmed cases. However, in terms of the confirmed infection rate, London was quickly overtaken by other regions; by 13 May, COVID-19 rates ranged from 128 per 100,000 in the South West to 348 per 100,000 in the North East,^1^ and hospital admission rates remained highest in the northern regions,^5^ where baseline health tends to be poorer.^6^ In addition to the impact of the virus itself, a second epidemic emerged as a result of the public health response. Many of the excess deaths during the pandemic are not associated with COVID-19 infection,^7^ and although this may be partly attributable to underdiagnosis, it raises the possibility that creating emergency capacity in primary and secondary care has compromised essential care. Patients have also avoided healthcare providers out of concern for service pressures or fear of infection, leading to delay of treatment and increases in out-of-hospital events such as cardiac arrest.^8 9^

If these non-COVID-19 excess deaths are also socially and geographically patterned, the consequences for health inequalities could be severe. To begin to address this issue, in this study we aimed to quantify excess mortality in England and Wales during the pandemic, as an aggregate and by region and age group, for all causes and for non-COVID-19 associated deaths.

## Methods

### Data

Weekly mortality data for the whole of England and Wales were obtained from the Office of National Statistics (ONS), covering the period from 1 Jan 2010 to 8 May 2020.^10^ All-cause deaths and deaths where the underlying cause was respiratory disease were available for England-Wales as an aggregate, but also by region (former Strategic Health Authority) and age group (under 1, 1-14, 15-44, 45-66, 65-74, 75-84 and 85 or over) for males and females. Respiratory disease deaths excluded deaths at age under 28 days and were obtained using ICD-10 codes J00-J99. For 2020 data, deaths where COVID-19 was mentioned in the death certificate were also reported (ICD10 U07.1 and U07.2), as England-Wales aggregates but also by region and age group. However, respiratory and COVID-19 deaths could be double-counted. Regional counts do not include residents outside England-Wales or those records where the place of residence is missing, while age group counts do not include deaths where age was missing.

Underlying cause of death, as reported in England and Wales, is compatible with WHO recommendations using ICD rules (with small changes in 2011 and 2014), and is obtained from the death certificate. The underlying cause of death is defined by the WHO as the disease or injury that initiated the series of events that led directly to death, or the circumstances of the accident or violence that produced the fatal injury.^11^ A condition mentioned in the death certificate may be the main reason or a contributory reason to the cause of death.

The study used aggregated national mortality data reported by the Office of National Statistics, and ethical approval was not needed.

### Analyses

Data were imported, cleaned and formatted as a time series in Stata v16. Death counts, overall and by stratum of interest were plotted and the same analysis model was used for each. For the excess mortality modelling, data from week 9 in 2020 (22-28 February), two weeks before the first official COVID-19 related deaths (7-13 March) were set to missing. Mean-dispersion negative binomial regressions were used to model death counts, with the fixed-effect predictors being week (as categorical, to account for seasonality) and time (as continuous, to account for a potential slope). For each model and stratum, the natural logarithm of the annual population estimate was used as offset. We selected negative binomial regression models over Poisson models because of the high variation in the outcome variable. For weeks 11 (7 to 13 March) to 19 (2 to 8 May), the exponentiated linear prediction from each model (and its 95% confidence intervals) were subtracted from: 1) all-cause deaths; and 2) all-cause deaths minus deaths where COVID-19 was mentioned. Excess deaths estimates, in each stratum and overall, were summed across all weeks, with a pooled estimate for the standard error used to obtain the 95% confidence intervals.

## Results

### All-cause excess mortality

During the 9-week period from 7 March 2020 to 8 May 2020, overall excess deaths for England and Wales were 47,243 (95%CI: 46,671 to 47,815) (Table 1). This total includes negative values at the start of the study period, when excess deaths were lower than predicted from historical trends, continuing a pattern of relatively low mortality rates in the first 12 weeks of 2020 (Figure 1). London had the greatest number of excess deaths at 9,219 (95%CI: 9,185 to 9,254), followed by the North West with 6,951 (95%CI: 6,903 to 6,999) and the South East with 6,656 (95%CI: 6,588 to 6,723). The least affected regions were Wales with 1,757 (95%CI: 1,730 to 1,784) and the North East with 2,239 deaths (95%CI: 2,209 to 2,270). The age-groups with the largest numbers of excess deaths were 85 or over with 20,173 (95%CI: 19,975 to 20,370) and 75-84 with 14,414 (95%CI: 14,304 to 14,524). We estimated 7,021 (95%CI: 6,963 to 7,078) excess all-cause deaths for people aged 65-74 and 4,811 (95%CI: 4,768 to 4,853) for people aged 45-64.

**Table 1:**
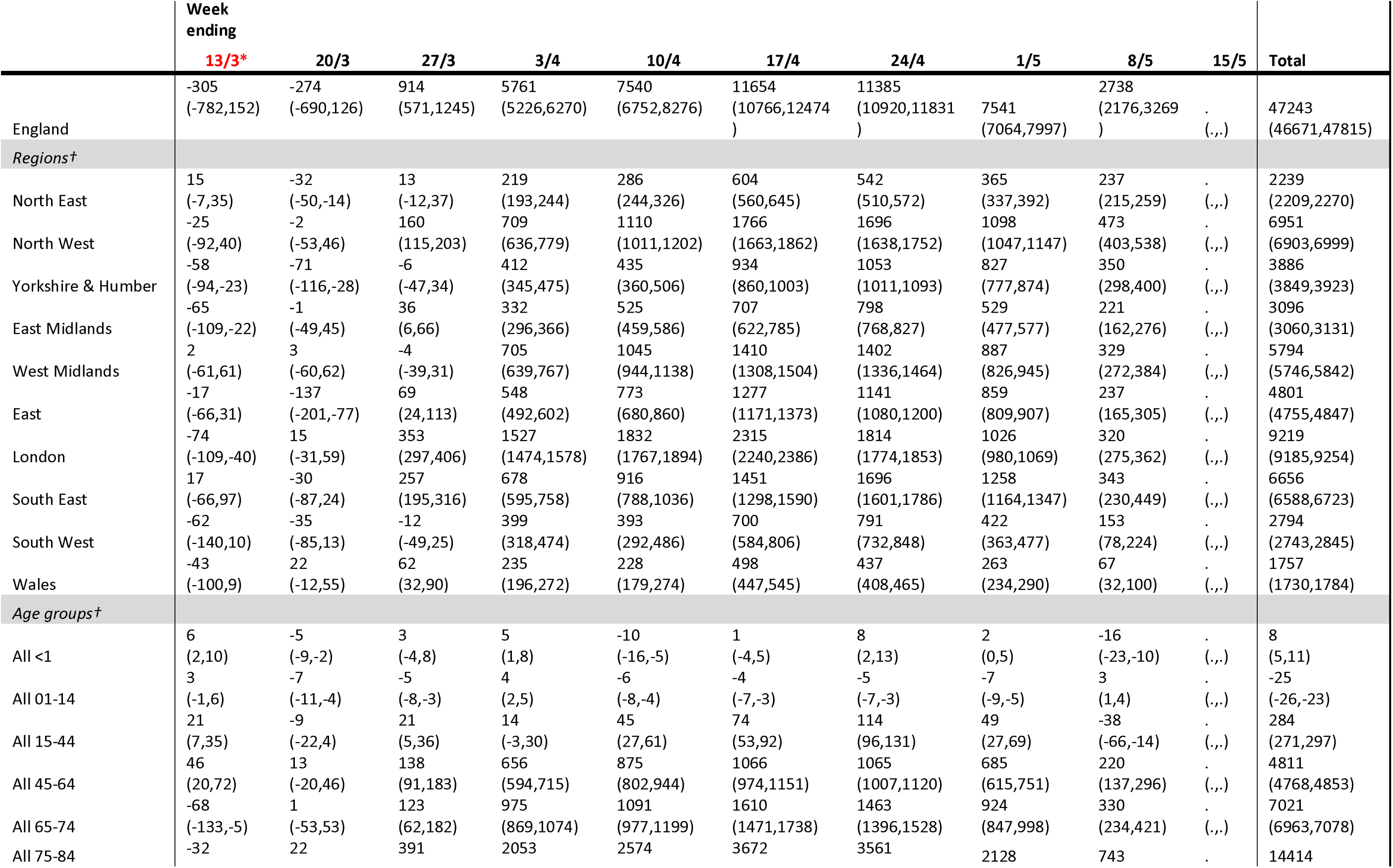
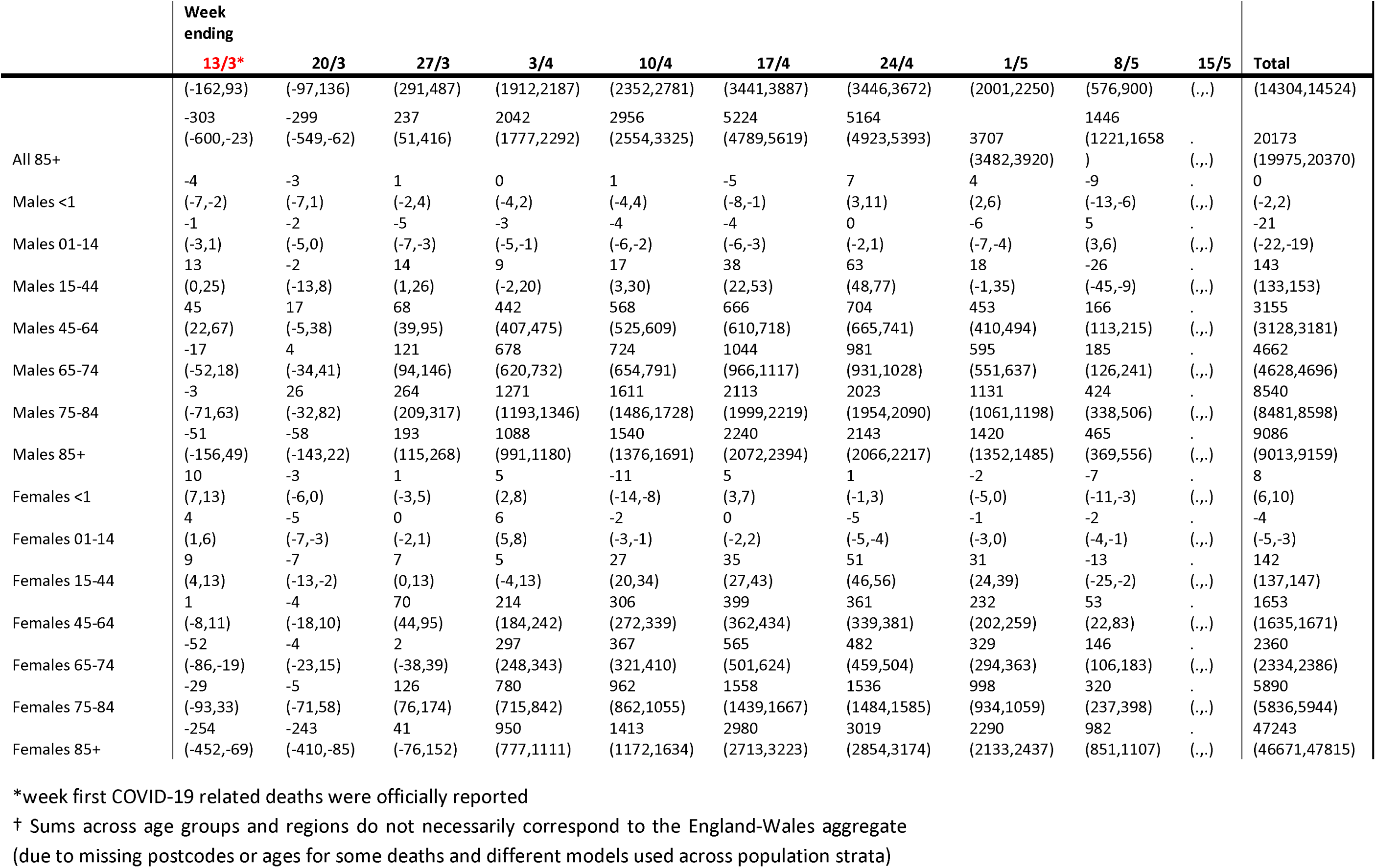
Excess all-cause deaths from negative binomial regression model, 22/2/2020 to 8/5/2020

**Figure 1:**
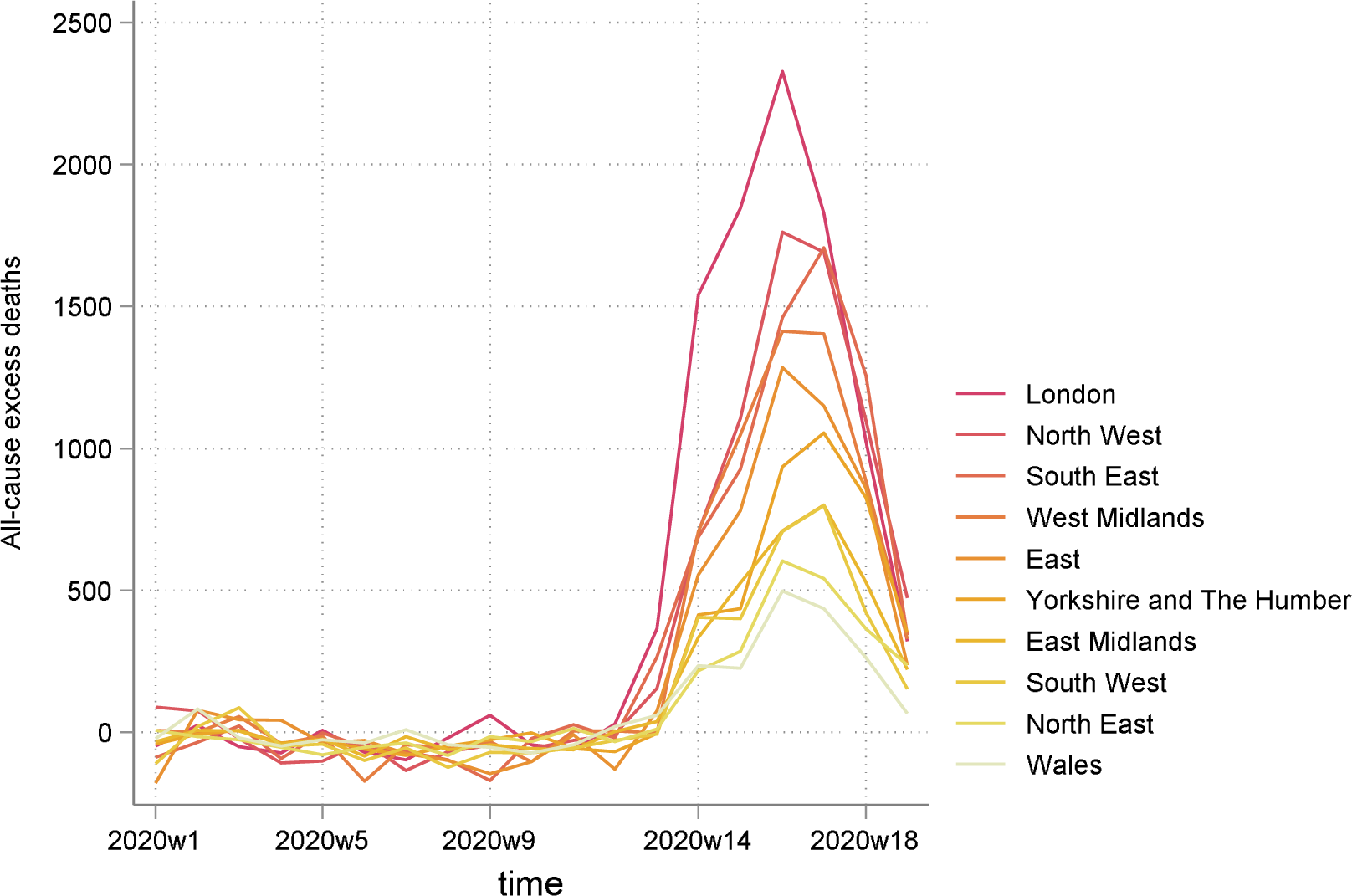
All-cause excess mortality by region, ranked by highest to lowest total.

All-cause excess mortality per 100,000 population is presented in Table 2, with the average across England and Wales at 79 (95%CI: 78 to 80). The largest mortality ratios were observed in London (102; 95%CI: 102 to 103), the West Midlands (97; 95%CI: 96 to 98) and the North West (94; 95%CI: 93 to 95), and the lowest in the South West (49; 95%CI: 49 to 50) and Wales (55; 95%CI: 55 to 56) (Figure 2). The mortality ratios for people aged 85 or over, 75-84, 65-74 and 45-64 were 1,331 (95%CI: 1,318 to 1,344), 388 (95%CI: 385 to 391), 118 (95%CI: 117 to 117) and 31 (95%CI: 31 to 32), respectively.

**Table 2:**
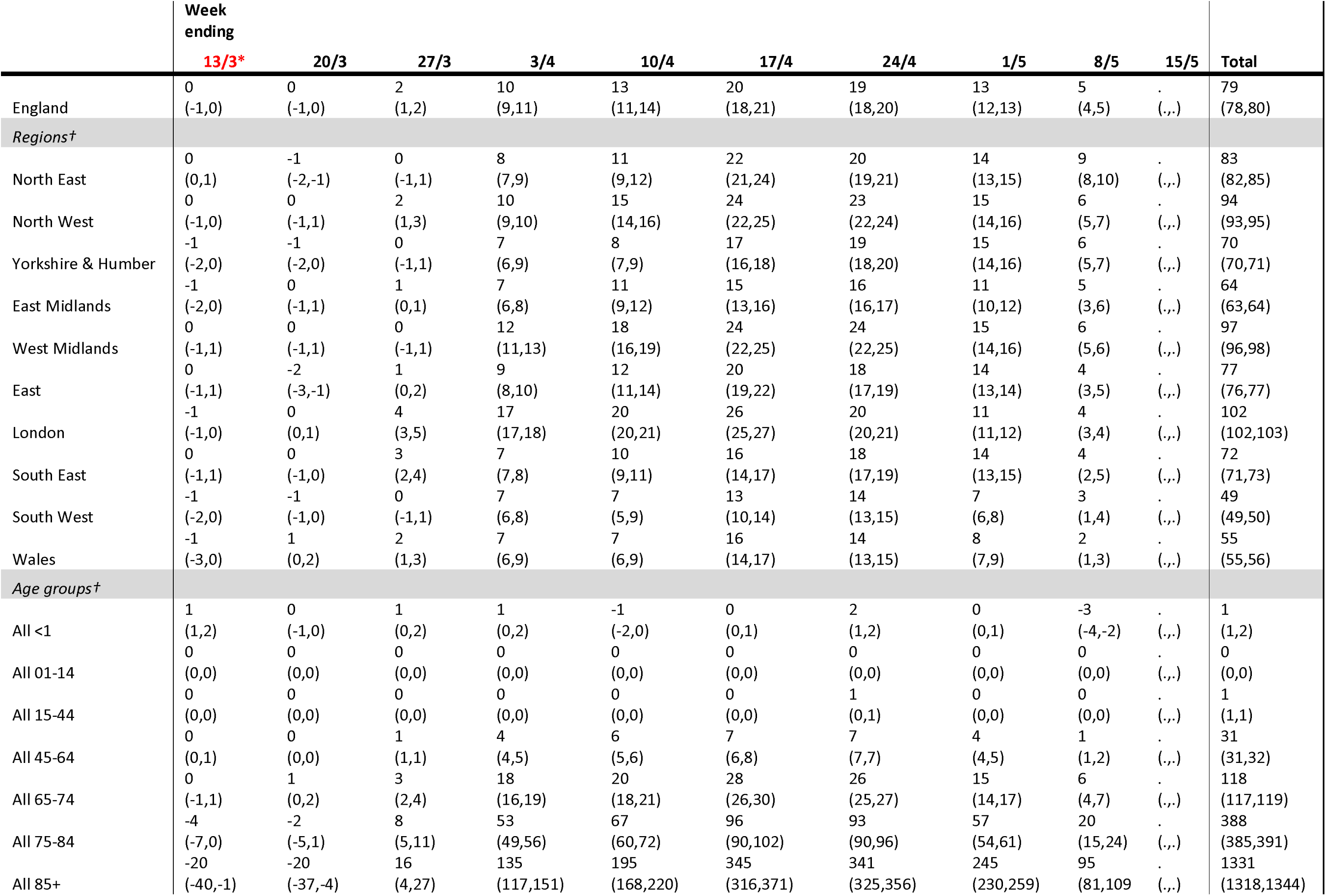
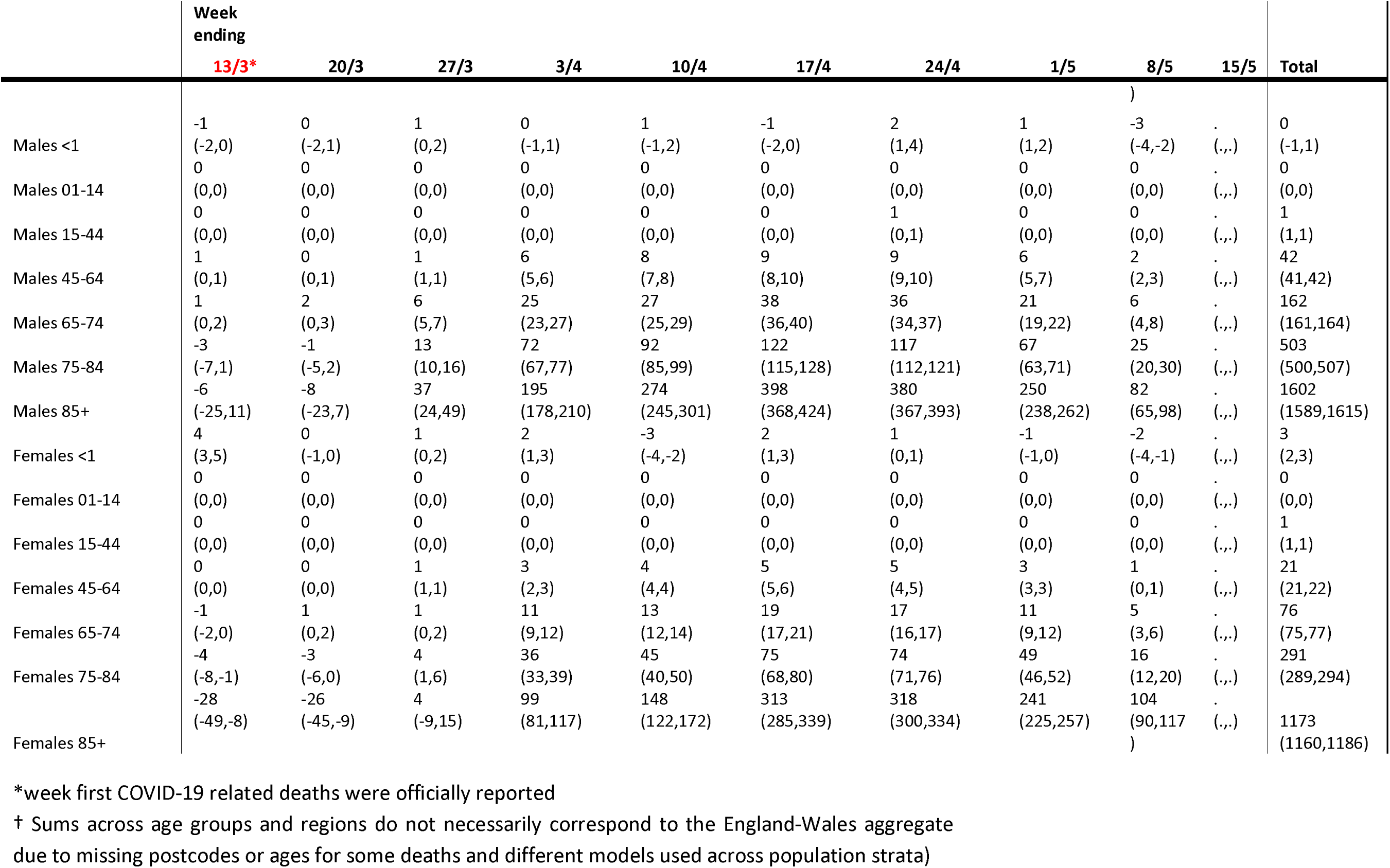
Excess all-cause deaths per 100,000 population from negative binomial regression model, 22/2/2020 to 8/5/2020

**Figure 2:**
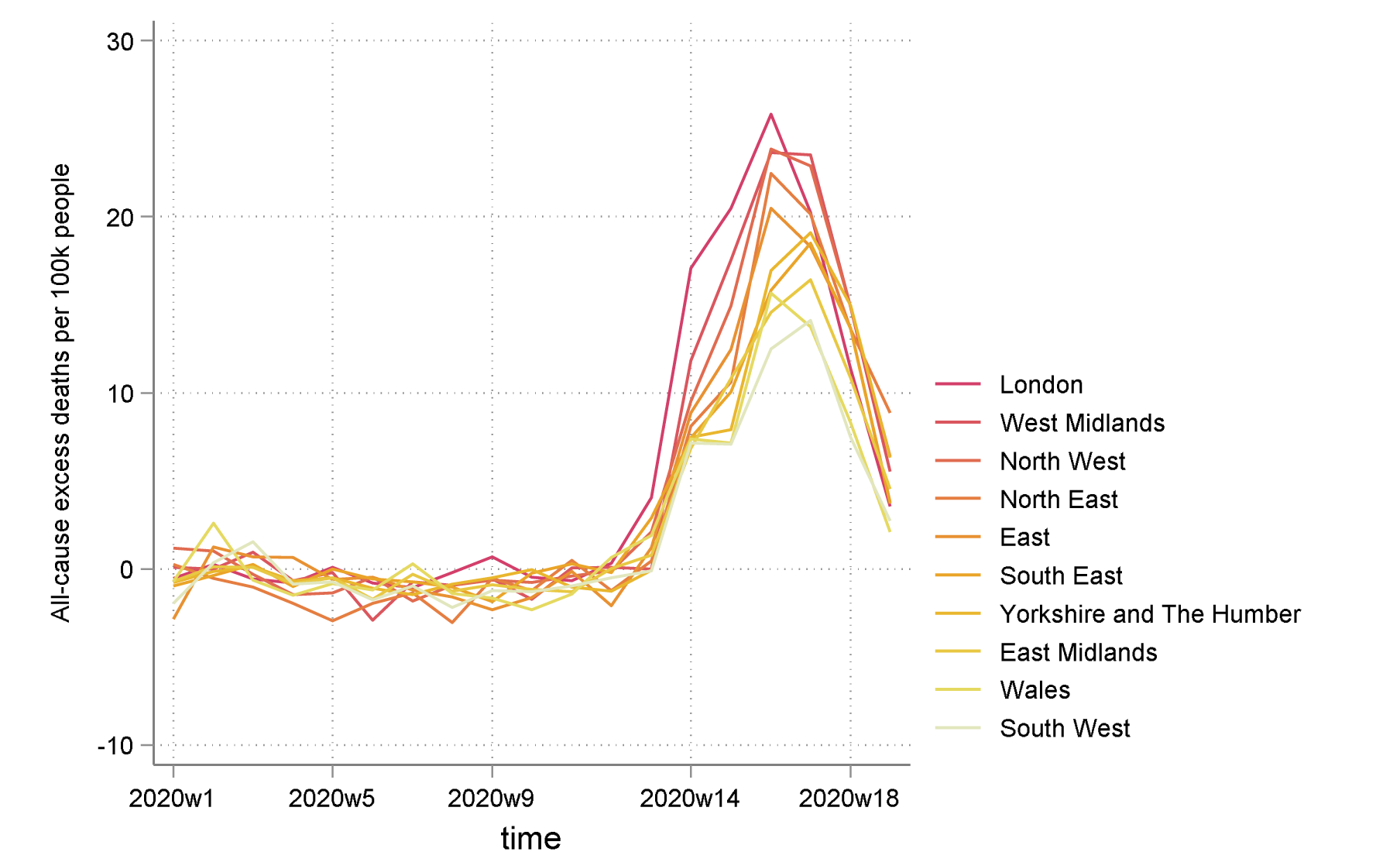
All-cause excess mortality per 100,000 population by region, ranked by highest to lowest total.

Excess all-cause deaths were estimated for all male and female age groups, and were higher for males for all age groups from age 15, with the exception of the 85+ age group. We estimated 9,086 (95%CI: 9,013 to 9,159) excess all-cause deaths for males aged 85 or over, compared to 11,123 for females (95%CI: 10,997 to 11,250). However, males aged 75-84 were estimated to have had 8,540 deaths (95%CI: 8,481 to 8,598), compared to 5,890 (95%CI: 5,836 to 5,944) for females, and males aged 65-74 were estimated to have had 4,662 deaths (95%CI: 4,628 to 4,696), compared to 2,360 (95%CI: 2,334 to 2,386) for females.

Mortality ratios per 100,000 population more clearly demonstrated the higher excess deaths in males. For males aged 85 or over, 75-84, 65-74 and 45-64, these were estimated at 1,602 (95%CI: 1,589 to 1,615), 503 (95%CI: 500 to 507), 162 (95%CI: 161 to 164) and 42 (95%CI: 41 to 42). Female estimates for the respective age groups were 1,173 (95%CI: 1,160 to 1,186), 291 (95%CI: 289 to 294), 76 (95%CI: 75 to 77) and 21 (95%CI: 21 to 22).

### All-cause excess mortality not attributed to COVID-19

From 7 March 2020 to 8 May 2020, overall excess deaths for England and Wales, excluding cases where COVID-19 was mentioned in the death certificate, were 9,948 (95%CI: 9,376 to 10,520) (Table 3). It is important to note that until the week ending 1 May 2020, the total was 11,141 (95%CI: 10,567 to 11,714), but there was a large drop into negative values in the last week of analysis (2 May to 8 May 2020). London was again the worst hit region in terms of number of excess deaths with 1,814 (95%CI: 1,780 to 1,849), followed by the South East with 1,570 (95%CI: 1,502 to 1,637) and the West Midlands with 1,531 (95%CI: 1,483 to 1,579) (Figure 3). The least affected regions were again Wales with −18 (95%CI: −45 to 9) and the North East with 320 (95%CI: 290 to 351). The age-groups with the largest numbers of excess deaths were 85 or over with 5,124 (95%CI: 4,926 to 5,321) and 75-84 with 2,207 (95%CI: 2,097 to 2,317). Estimated excess deaths for people aged 65-74 and 45-64 were 1,249 (95%CI: 1,191 to 1,306) and 969 (95%CI: 926 to 1,011), respectively.

**Table 3:** Excess all-cause deaths, minus those with documented COVID-19 infection, from negative binomial regression model, 22/2/2020 to 8/5/2020

|  | Week<br>ending |  |  |  |  |  |  |  |  |  |  |
| --- | --- | --- | --- | --- | --- | --- | --- | --- | --- | --- | --- |
|  | 13/3* | 20/3 | 27/3 | 3/4 | 10/4 | 17/4 | 24/4 | 1/5 | 8/5 | 15/5 | Total |
| England | -310<br>(-787,147) | -377<br>(-793,23) | 375<br>(32,706) | 2286<br>(1751,2795) | 1327<br>(539,2063) | 2896<br>(2008,3716) | 3148<br>(2683,3594) | 1506<br>(1029,1962) | -1192<br>(-1754,-661) | . | 9948<br>(9376,10520) |
| *Regions†* |  |  |  |  |  |  |  |  |  |  |  |
| North East | 15<br>(-7,35) | -33<br>(-50,-14) | -2<br>(-12,37) | 85<br>(193,244) | -2<br>(244,326) | 129<br>(560,645) | 123<br>(510,572) | 49<br>(337,392) | -34<br>(215,259) | . | 320<br>(290,351) |
| North West | -26<br>(-92,40) | -14<br>(-53,46) | 100<br>(115,203) | 291<br>(636,779) | 201<br>(1011,1202) | 416<br>(1663,1862) | 489<br>(1638,1752) | 188<br>(1047,1147) | -124<br>(403,538) | . | 1487<br>(1439,1535) |
| Yorkshire & Humber | -58<br>(-94,-23) | -76<br>(-116,-28) | -18<br>(-47,34) | 238<br>(345,475) | 61<br>(360,506) | 244<br>(860,1003) | 298<br>(1011,1093) | 180<br>(777,874) | -82<br>(298,400) | . | 797<br>(760,834) |
| East Midlands | -65<br>(-109,-22) | -4<br>(-49,45) | 12<br>(6,66) | 147<br>(296,366) | 101<br>(459,586) | 149<br>(622,785) | 236<br>(768,827) | 91<br>(477,577) | -113<br>(162,276) | . | 568<br>(532,603) |
| West Midlands | 0<br>(-61,61) | -11<br>(-60,62) | -71<br>(-39,31) | 305<br>(639,767) | 238<br>(944,1138) | 411<br>(1308,1504) | 522<br>(1336,1464) | 236<br>(826,945) | -114<br>(272,384) | . | 1531<br>(1483,1579) |
| East | -17<br>(-66,31) | -140<br>(-201,-77) | 57<br>(24,113) | 265<br>(492,602) | 209<br>(680,860) | 442<br>(1171,1373) | 327<br>(1080,1200) | 228<br>(809,907) | -136<br>(165,305) | . | 1286<br>(1240,1332) |
| London | -74<br>(-109,-40) | -29<br>(-31,59) | 116<br>(297,406) | 357<br>(1474,1578) | 326<br>(1767,1894) | 497<br>(2240,2386) | 408<br>(1774,1853) | 241<br>(980,1069) | -119<br>(275,362) | . | 1814<br>(1780,1849) |
| South East | 15<br>(-66,97) | -47<br>(-87,24) | 188<br>(195,316) | 267<br>(595,758) | 187<br>(788,1036) | 342<br>(1298,1590) | 464<br>(1601,1786) | 292<br>(1164,1347) | -208<br>(230,449) | . | 1570<br>(1502,1637) |
| South West | -62<br>(-140,10) | -36<br>(-85,13) | -31<br>(-49,25) | 244<br>(318,474) | 95<br>(292,486) | 199<br>(584,806) | 260<br>(732,848) | 18<br>(363,477) | -123<br>(78,224) | . | 609<br>(558,660) |
| Wales | -43<br>(-100,9) | 20<br>(-12,55) | 41<br>(32,90) | 101<br>(196,272) | -76<br>(179,274) | 89<br>(447,545) | 24<br>(408,465) | -18<br>(234,290) | -144<br>(32,100) | . | -18<br>(-45,9) |
| *Age groups†* |  |  |  |  |  |  |  |  |  |  |  |
| All <1 | 6<br>(2,10) | -5<br>(-9,-2) | 3<br>(-4,8) | 5<br>(1,8) | -10<br>(-16,-5) | 1<br>(-4,5) | 8<br>(2,13) | 2<br>(0,5) | -17<br>(-24,-11) | . | 7<br>(4,10) |
| All 01-14 | 3<br>(-1,6) | -7<br>(-11,-4) | -5<br>(-8,-3) | 4<br>(2,5) | -6<br>(-8,-4) | -6<br>(-9,-5) | -5<br>(-7,-3) | -7<br>(-9,-5) | 3<br>(1,4) | . | -27<br>(-28,-25) |
| All 15-44 | 21<br>(7,35) | -10<br>(-23,3) | 14<br>(-2,29) | -35<br>(-52,-19) | -33<br>(-51,-17) | -30<br>(-51,-12) | 5<br>(-13,22) | -5<br>(-27,15) | -78<br>(-106,-54) | . | -138<br>(-151,-125) |
| All 45-64 | 45<br>(19,71) | 6<br>(-27,39) | 63<br>(16,108) | 210<br>(148,269) | 70<br>(-3,139) | 46<br>(-46,131) | 182<br>(124,237) | 130<br>(60,196) | -97<br>(-180,-21) | . | 969<br>(926,1011) |
| All 65-74 | -69<br>(-134,-6) | -22<br>(-76,30) | 10<br>(-51,69) | 274<br>(168,373) | -128<br>(-242,-20) | 84<br>(-55,212) | 169<br>(102,234) | 34<br>(-43,108) | -156<br>(-252,-65) | . | 1249<br>(1191,1306) |
| All 75-84 | -35<br>(-165,90) | -13<br>(-132,101) | 184<br>(84,280) | 702<br>(561,836) | 109<br>(-113,316) | 581<br>(350,796) | 685<br>(570,796) | 84<br>(-43,206) | -493<br>(-660,-336) | . | 2207<br>(2097,2317) |
| All 85+ | -303<br>(-600,-23) | -346<br>(-596,-109) | 32<br>(-154,211) | 788<br>(523,1038) | 636<br>(234,1005) | 1471<br>(1036,1866) | 1323<br>(1082,1552) | 574<br>(349,787) | -404<br>(-629,-192) | . | 5124<br>(4926,5321) |
|  | 13/3* | 20/3 | 27/3 | 3/4 | 10/4 | 17/4 | 24/4 | 1/5 | 8/5 | 15/5 | Total |
|  | -4 | -3 | 1 | 0 | 1 | -5 | 7 | 4 | -10 | . | -1 |
| Males <1 | (-7,-2) | (-7,1) | (-2,4) | (-4,2) | (-4,4) | (-8,-1) | (3,11) | (2,6) | (-14,-7) | (.,.) | (-3,1) |
|  | -1 | -2 | -5 | -3 | -4 | -4 | 0 | -6 | 5 | . | -21 |
| Males 01-14 | (-3,1) | (-5,0) | (-7,-3) | (-5,-1) | (-6,-2) | (-6,-3) | (-2,1) | (-7,-4) | (3,6) | (.,.) | (-22,-19) |
|  | 13 | -3 | 12 | -16 | -32 | -24 | -5 | -17 | -46 | . | -113 |
| Males 15-44 | (0,25) | (-14,7) | (-1,24) | (-27,-5) | (-46,-19) | (-40,-9) | (-20,9) | (-36,0) | (-65,-29) | (.,.) | (-123,-103) |
|  | 44 | 14 | 36 | 158 | 42 | -3 | 117 | 83 | -47 | . | 614 |
| Males 45-64 | (21,66) | (-8,35) | (7,63) | (123,191) | (-1,83) | (-59,49) | (78,154) | (40,124) | (-100,2) | (.,.) | (587,640) |
|  | -17 | -4 | 82 | 212 | -68 | 38 | 156 | 13 | -115 | . | 863 |
| Males 65-74 | (-52,18) | (-42,33) | (55,107) | (154,266) | (-138,-1) | (-40,111) | (106,203) | (-31,55) | (-174,-59) | (.,.) | (829,897) |
|  | -4 | 11 | 186 | 421 | 69 | 267 | 364 | 38 | -285 | . | 1214 |
| Males 75-84 | (-72,62) | (-47,67) | (131,239) | (343,496) | (-56,186) | (153,373) | (295,431) | (-32,105) | (-371,-203) | (.,.) | (1155,1272) |
|  | -51 | -77 | 102 | 431 | 288 | 433 | 427 | 102 | -284 | . | 1910 |
| Males 85+ | (-156,49) | (-162,3) | (24,177) | (334,523) | (124,439) | (265,587) | (350,501) | (34,167) | (-380,-193) | (.,.) | (1837,1983) |
|  | 10 | -3 | 1 | 5 | -11 | 5 | 1 | -2 | -7 | . | 8 |
| Females <1 | (7,13) | (-6,0) | (-3,5) | (2,8) | (-14,-8) | (3,7) | (-1,3) | (-5,0) | (-11,-3) | (.,.) | (6,10) |
|  | 4 | -5 | 0 | 6 | -2 | -2 | -5 | -1 | -2 | . | -6 |
| Females 01-14 | (1,6) | (-7,-3) | (-2,1) | (5,8) | (-3,-1) | (-4,0) | (-5,-4) | (-3,0) | (-4,-1) | (.,.) | (-7,-5) |
|  | 9 | -7 | 1 | -17 | -1 | -6 | 10 | 12 | -33 | . | -24 |
| Females 15-44 | (4,13) | (-13,-2) | (-6,7) | (-26,-9) | (-8,6) | (-14,2) | (5,15) | (5,20) | (-45,-22) | (.,.) | (-29,-19) |
|  | 1 | -8 | 27 | 55 | 29 | 57 | 76 | 53 | -51 | . | 352 |
| Females 45-64 | (-8,11) | (-22,6) | (1,52) | (25,83) | (-5,62) | (20,92) | (54,96) | (23,80) | (-82,-21) | (.,.) | (334,370) |
|  | -53 | -20 | -69 | 70 | -43 | 56 | 30 | 36 | -40 | . | 387 |
| Females 65-74 | (-87,-20) | (-39,-1) | (-109,-32) | (21,116) | (-89,0) | (-8,115) | (7,52) | (1,70) | (-80,-3) | (.,.) | (361,413) |
|  | -31 | -24 | -1 | 293 | 71 | 350 | 356 | 84 | -207 | . | 1009 |
| Females 75-84 | (-95,31) | (-90,39) | (-51,47) | (228,355) | (-29,164) | (231,459) | (304,405) | (20,145) | (-290,-129) | (.,.) | (955,1063) |
|  | -254 | -271 | -71 | 370 | 369 | 1077 | 957 | 541 | -119 | . | 3250 |
| Females 85+ | (-452,-69) | (-438,-113) | (-188,40) | (197,531) | (128,590) | (810,1320) | (792,1112) | (384,688) | (-250,6) | (.,.) | (3124,3377) |

\*week first COVID-19 related deaths were officially reported
† Sums across age groups and regions do not necessarily correspond to the England-Wales aggregate due to missing postcodes or ages for some deaths and different models used across population strata)

**Figure 3:**
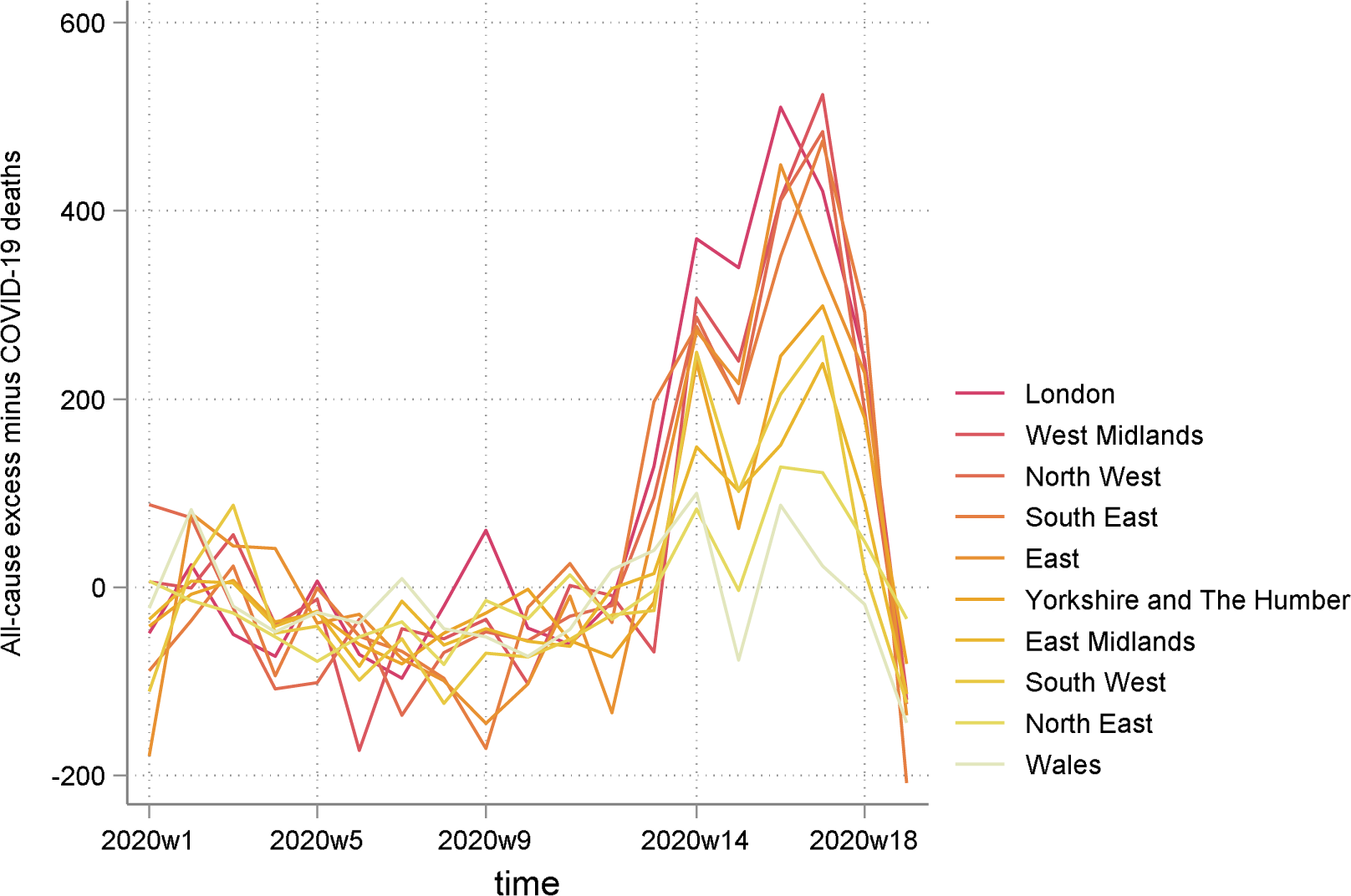
All-cause minus COVID-19 related excess mortality by region, ranked by highest to lowest total.

Non-COVID-19 excess mortality per 100,000 population is presented in Table 4, with the average across England and Wales at 17 (95%CI: 16 to 18). The largest mortality ratios were in the West Midlands (26; 95%CI: 25 to 26), followed by the East of England (21; 95%CI: 20 to 21), the North West (20; 95%CI: 19 to 21) and London (20; 95%CI: 20 to 21) (Figure 4). Ratios for people aged 85 or over, 75-84, 65-74 and 45-64 were 338 (95%CI: 325 to 351), 59 (95%CI: 56 to 62), 21 (95%CI: 20 to 22) and 6 (95%CI: 6 to 7), respectively.

**Table 4:** Excess all-cause deaths, minus those with documented COVID-19 infection, per 100,000 population, from negative binomial regression model, 22/2/2020 to 8/5/2020

|  | Week<br>ending |  |  |  |  |  |  |  |  |  |  |
| --- | --- | --- | --- | --- | --- | --- | --- | --- | --- | --- | --- |
|  | 13/3* | 20/3 | 27/3 | 3/4 | 10/4 | 17/4 | 24/4 | 1/5 | 8/5 | 15/5 | Total |
|  | 0 | -1 | 1 | 4 | 2 | 5 | 5 | 3 | -2 | . | 17 |
| England | (-1,0) | (-1,0) | (0,1) | (3,5) | (1,4) | (3,6) | (5,6) | (2,3) | (-3,-1) | (.,.) | (16,18) |
| *Regions†* |  |  |  |  |  |  |  |  |  |  |  |
|  | -1 | -1 | 0 | -1 | 0 | 3 | 0 | 5 | 5 | . | 12 |
| North East | (-1,0) | (-2,0) | (0,1) | (-2,-1) | (-1,1) | (2,4) | (-2,1) | (3,6) | (3,6) | (.,.) | (11,13) |
|  | -1 | -1 | 0 | 0 | 1 | 4 | 3 | 6 | 7 | . | 20 |
| North West | (-1,0) | (-2,0) | (-1,0) | (-1,0) | (1,2) | (3,5) | (1,4) | (4,7) | (6,7) | (.,.) | (19,21) |
|  | 0 | 0 | -1 | -1 | 0 | 4 | 1 | 4 | 5 | . | 14 |
| Yorkshire & Humber | (-1,0) | (-1,1) | (-2,0) | (-2,-1) | (-1,0) | (3,5) | (0,2) | (3,6) | (5,6) | (.,.) | (14,15) |
|  | -1 | -1 | -1 | 0 | 0 | 3 | 2 | 3 | 5 | . | 12 |
| East Midlands | (-2,0) | (-2,0) | (-2,0) | (-1,1) | (0,1) | (2,4) | (1,3) | (1,5) | (4,5) | (.,.) | (11,12) |
|  | -1 | -2 | 0 | 0 | -1 | 5 | 4 | 7 | 9 | . | 26 |
| West Midlands | (-1,0) | (-3,-1) | (-1,1) | (-1,1) | (-2,-1) | (4,6) | (2,6) | (5,8) | (8,10) | (.,.) | (25,26) |
|  | -2 | -2 | 0 | -2 | 1 | 4 | 3 | 7 | 5 | . | 21 |
| East | (-3,-1) | (-3,-1) | (-1,1) | (-3,-1) | (0,2) | (3,5) | (2,5) | (5,9) | (4,6) | (.,.) | (20,21) |
|  | 1 | 0 | -1 | 0 | 1 | 4 | 4 | 6 | 5 | . | 20 |
| London | (0,1) | (-1,0) | (-1,0) | (-1,0) | (1,2) | (4,5) | (3,4) | (5,6) | (4,5) | (.,.) | (20,21) |
|  | -2 | 0 | 0 | 0 | 2 | 3 | 2 | 4 | 5 | . | 17 |
| South East | (-2,-1) | (-1,1) | (-1,1) | (-1,0) | (1,3) | (2,4) | (1,3) | (2,5) | (4,6) | (.,.) | (16,18) |
|  | -1 | -1 | -1 | -1 | 0 | 4 | 2 | 4 | 5 | . | 11 |
| South West | (-2,0) | (-3,0) | (-2,0) | (-1,0) | (-1,0) | (3,6) | (0,3) | (2,5) | (4,6) | (.,.) | (10,12) |
|  | -2 | -2 | -1 | 1 | 1 | 3 | -2 | 3 | 1 | . | -1 |
| Wales | (-3,-1) | (-4,-1) | (-3,0) | (-1,2) | (0,2) | (2,4) | (-4,-1) | (1,4) | (0,2) | (.,.) | (-1,0) |
| *Age groups†* |  |  |  |  |  |  |  |  |  |  |  |
|  | 1 | 0 | 1 | 1 | -1 | 0 | 2 | 0 | -3 | . | 1 |
| All <1 | (1,2) | (-1,0) | (0,2) | (0,2) | (-2,0) | (0,1) | (1,2) | (0,1) | (-4,-2) | (.,.) | (1,2) |
|  | 0 | 0 | 0 | 0 | 0 | 0 | 0 | 0 | 0 | . | 0 |
| All 01-14 | (0,0) | (0,0) | (0,0) | (0,0) | (0,0) | (0,0) | (0,0) | (0,0) | (0,0) | (.,.) | (0,0) |
|  | 0 | 0 | 0 | 0 | 0 | 0 | 0 | 0 | 0 | . | -1 |
| All 15-44 | (0,0) | (0,0) | (0,0) | (0,0) | (0,0) | (0,0) | (0,0) | (0,0) | (0,0) | (.,.) | (-1,-1) |
|  | 0 | 0 | 1 | 2 | 1 | 1 | 2 | 1 | -1 | . | 6 |
| All 45-64 | (0,1) | (0,0) | (0,1) | (1,2) | (0,1) | (0,1) | (1,2) | (1,2) | (-1,0) | (.,.) | (6,7) |
|  | 0 | 1 | 2 | 7 | 1 | 4 | 6 | 2 | -3 | . | 21 |
| All 65-74 | (-1,1) | (0,2) | (1,3) | (6,9) | (-1,3) | (2,6) | (5,7) | (1,3) | (-4,-1) | (.,.) | (20,22) |
|  | -4 | -3 | 3 | 19 | 7 | 20 | 23 | 7 | -13 | . | 59 |
| All 75-84 | (-8,0) | (-6,0) | (0,6) | (16,23) | (1,13) | (13,26) | (19,26) | (4,10) | (-18,-9) | (.,.) | (56,62) |
|  | -20 | -23 | 3 | 58 | 58 | 119 | 109 | 60 | -27 | . | 338 |
| All 85+ | (-40,-1) | (-40,-7) | (-9,15) | (40,75) | (31,82) | (91,146) | (93,124) | (45,74) | (-41,-13) | (.,.) | (325,351) |
|  | 13/3* | 20/3 | 27/3 | 3/4 | 10/4 | 17/4 | 24/4 | 1/5 | 8/5 | 15/5 | Total |
|  | -1 | 0 | 1 | 0 | 1 | -1 | 2 | 1 | -3 | . | 0 |
| Males <1 | (-2,0) | (-2,1) | (0,2) | (-1,1) | (-1,2) | (-2,0) | (1,4) | (1,2) | (-4,-2) | (.,.) | (-1,0) |
|  | 0 | 0 | 0 | 0 | 0 | 0 | 0 | 0 | 0 | . | 0 |
| Males 01-14 | (0,0) | (0,0) | (0,0) | (0,0) | (0,0) | (0,0) | (0,0) | (0,0) | (0,0) | (.,.) | (0,0) |
|  | 0 | 0 | 0 | 0 | 0 | 0 | 0 | 0 | 0 | . | -1 |
| Males 15-44 | (0,0) | (0,0) | (0,0) | (0,0) | (0,0) | (0,0) | (0,0) | (0,0) | (-1,0) | (.,.) | (-1,-1) |
|  | 1 | 0 | 0 | 2 | 1 | 0 | 2 | 1 | -1 | . | 8 |
| Males 45-64 | (0,1) | (0,1) | (0,1) | (2,3) | (1,2) | (0,1) | (2,3) | (1,2) | (-1,0) | (.,.) | (8,8) |
|  | 1 | 1 | 3 | 11 | 2 | 5 | 9 | 2 | -4 | . | 30 |
| Males 65-74 | (0,2) | (0,3) | (3,4) | (9,13) | (-1,4) | (2,7) | (7,10) | (1,4) | (-6,-2) | (.,.) | (29,31) |
|  | -3 | -2 | 6 | 25 | 9 | 21 | 26 | 7 | -17 | . | 72 |
| Males 75-84 | (-7,1) | (-6,1) | (3,9) | (21,30) | (1,16) | (14,27) | (22,30) | (3,11) | (-22,-12) | (.,.) | (68,75) |
|  | -6 | -12 | 19 | 84 | 71 | 102 | 95 | 35 | -50 | . | 337 |
| Males 85+ | (-25,11) | (-28,2) | (6,31) | (67,100) | (42,98) | (73,128) | (82,107) | (23,46) | (-67,-34) | (.,.) | (324,350) |
|  | 4 | 0 | 1 | 2 | -3 | 2 | 1 | -1 | -2 | . | 3 |
| Females <1 | (3,5) | (-1,0) | (0,2) | (1,3) | (-4,-2) | (1,3) | (0,1) | (-1,0) | (-4,-1) | (.,.) | (2,3) |
|  | 0 | 0 | 0 | 0 | 0 | 0 | 0 | 0 | 0 | . | 0 |
| Females 01-14 | (0,0) | (0,0) | (0,0) | (0,0) | (0,0) | (0,0) | (0,0) | (0,0) | (0,0) | (.,.) | (0,0) |
|  | 0 | 0 | 0 | 0 | 0 | 0 | 0 | 0 | 0 | . | 0 |
| Females 15-44 | (0,0) | (0,0) | (0,0) | (0,0) | (0,0) | (0,0) | (0,0) | (0,0) | (0,0) | (.,.) | (0,0) |
|  | 0 | 0 | 1 | 1 | 1 | 1 | 1 | 1 | -1 | . | 5 |
| Females 45-64 | (0,0) | (0,0) | (0,1) | (1,1) | (0,1) | (1,1) | (1,1) | (0,1) | (-1,0) | (.,.) | (4,5) |
|  | -1 | 1 | 0 | 4 | 1 | 4 | 3 | 2 | -1 | . | 12 |
| Females 65-74 | (-2,0) | (0,1) | (-1,1) | (2,5) | (-1,2) | (2,5) | (3,4) | (1,3) | (-3,0) | (.,.) | (12,13) |
|  | -4 | -3 | 1 | 14 | 6 | 19 | 20 | 7 | -10 | . | 50 |
| Females 75-84 | (-8,-1) | (-7,0) | (-2,3) | (11,18) | (1,10) | (13,25) | (17,23) | (4,10) | (-14,-6) | (.,.) | (47,53) |
|  | -28 | -28 | -6 | 43 | 50 | 131 | 119 | 75 | -13 | . | 343 |
| Females 85+ | (-49,-8) | (-47,-11) | (-18,6) | (24,60) | (24,74) | (102,156) | (101,135) | (59,91) | (-26,1) | (.,.) | (329,356) |

\*week first COVID-19 related deaths were officially reported
† Sums across age groups and regions do not necessarily correspond to the England-Wales aggregate due to missing postcodes or ages for some deaths and different models used across population strata)

**Figure 4:**
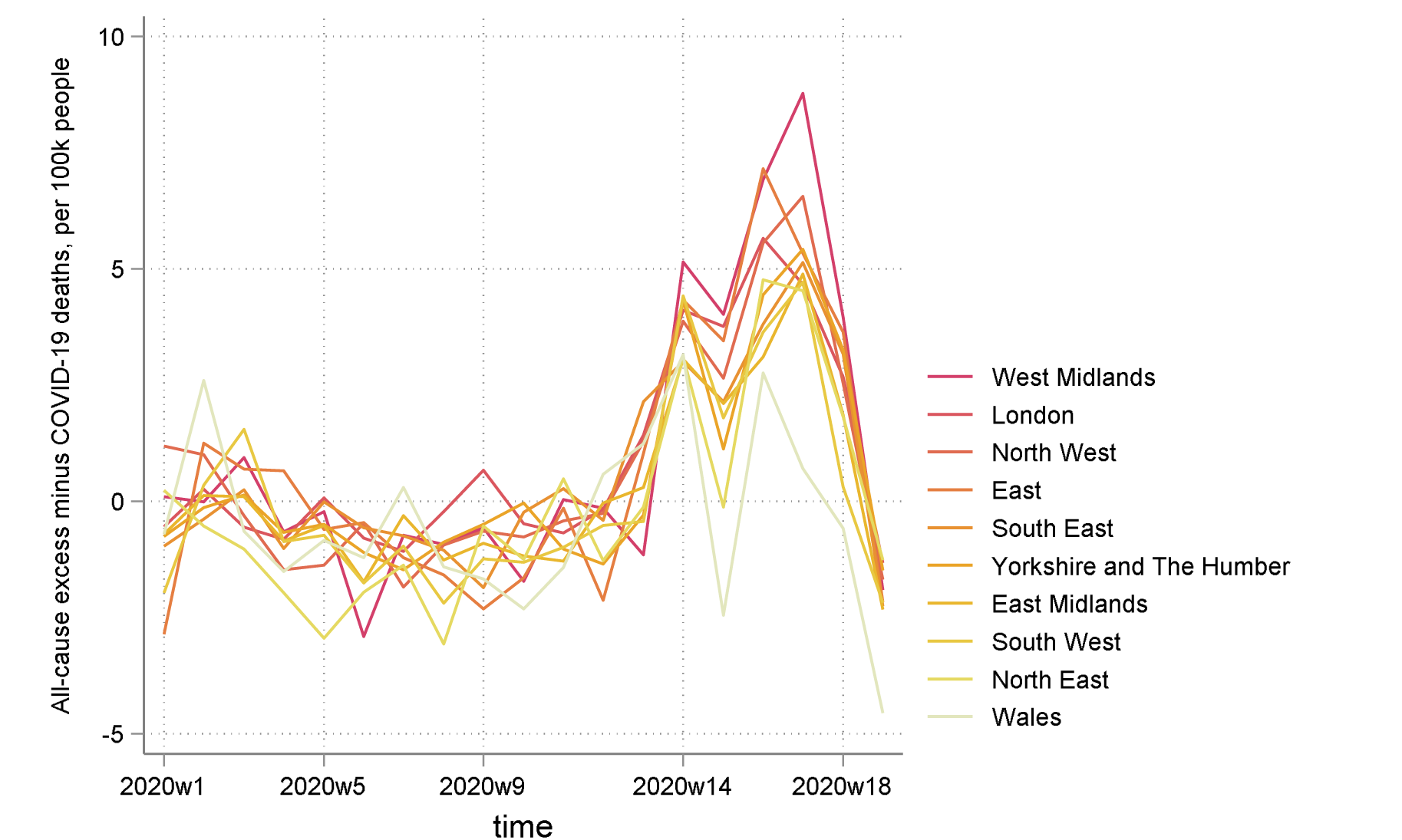
All-cause minus COVID-19 related excess mortality per 100,000 population by region, ranked by highest to lowest total.

Excess all-cause deaths, excluding deaths where COVID-19 was documented cause of death on the death certificate, were higher for males for all age groups from age 45, with the exception of the 85+ age group. We estimated 3,250 (95%CI: 3,124 to 3,377) excess deaths for females aged 85 or over, compared to 1,910 for males (95%CI: 1,837 to 1,983). However, males were estimated to have had 1,214 (95%CI: 1,155 to 1,272), 863 (95%CI: 829 to 897) and 614 (95%CI: 587 to 640) excess deaths for age groups 75-84, 65-74 and 45-64, respectively. By comparison, females were estimated to have had 1,009 (95%CI: 955 to 1,063), 387 (95%CI: 361 to 413) and 352 (95%CI: 334 to 370) excess deaths for age groups 75-84, 65-75 and 45-64, respectively. Males also had higher non-COVID-19 mortality ratios per 100,000 population, with 337 (95%CI: 324 to 350) for males aged 85 or over, 72 (95%CI: 68 to 75) for males aged 75-84, and 30 (95%CI: 29 to 31) for males aged 65-74. Female estimates for the respective age groups were 343 (95%CI: 329 to 356), 50 (95%CI: 47 to 53) and 12 (95%CI: 12 to 13).

### Supplementary files

Three dynamic supplementary files are also provided, produced in LaTeX for almost complete automation, which will be updated weekly. The first appendix reports the graphs presented in the paper, time-series of excess deaths by region, but also time series of excess deaths by age group and sex. The second appendix reports absolute numbers, the third reports ratios over 100,000 population, for each stratum of interest. For the overall England-Wales aggregate, each region and each age group (sex specific or not), we report the following time-series: mortality, mortality and predictions from the negative binomial regression model, all-cause excess deaths and all-cause excess deaths where COVID-19 is not mentioned in the death certificate. All these time series are reported from the 1st week of 2010, the 1st week of 2019 and the 1st week of 2020 (to allow for a more complete overview).

## Discussion

Our models, based on historical mortality trends, illustrate the scale of the COVID-19 pandemic and its direct and indirect impacts on population health. Over the first nine weeks, there were 47,243 excess deaths (95%CI: 46,617 to 47,815) in England and Wales, of which over a fifth (9,948, 95%CI: 9,376 to 10,520) were not directly attributed to COVID-19 infection. This likely underestimates the impact of the virus, as prior to the pandemic mortality rates for 2020 were relatively low, particularly compared with years with high influenza activity (2015 and 2018). Excess death rates started to increase after 20 March and peaked between 17 and 24 April, with the highest peaks (up to 26 deaths per 100,000) in London, the West Midlands and the North West and the lowest peaks in the East Midlands, Wales and the South West. Non-COVID-19 associated deaths followed similar general patterns. As has been previously reported, we observed that males had larger excess mortality rates than females across all age groups. However, female excess mortality rates excluding COVID-19 were higher in the 85+ age group, indicating a large undocumented impact of the virus on older females, which is likely to be both direct and indirect.

## Strengths and Limitations

This analysis of national data for England and Wales provides a comprehensive picture of excess deaths due to COVID-19, either directly or indirectly, based on diagnosis by clinicians closely involved with patient care. This allows for clinical judgment of the contribution of pandemic coronavirus to death, and does not only rely on a positive test for COVID-19 infection.

However, there are several important limitations. Although counts of deaths are likely to be accurate, cause of death information is less reliable, as it is reliant on accurate diagnosis and recording by clinicians dealing with exceptional circumstances. ONS reports of COVID-19 related deaths are based on clinical judgments, which in some cases will have been made without positive PCR or antibody testing. Although this confers some advantages compared to solely relying on the presence of a positive test, particularly when the sensitivity of these tests varies between 60-80%, unexpected deaths in elderly multimorbid patients or in those with respiratory symptoms may be misattributed to COVID-19. We controlled for annual population size, while our analyses are weekly, since changes in the population structure are small and gradual and not available on a weekly basis. Data for the latest weeks are provisional, subject to updates in later ONS releases, and the ONS reports the date of registration, rather than date of death, resulting in fluctuations in counts during certain periods (for example, over Easter). Finally, in assessing the unintended consequences of the public health response, it is important to note that we estimated net excess deaths. We do not know how many deaths have been avoided by adopting public health social isolation measures, either through reducing exposure to COVID-19 itself or through spill-over benefits such as reduced road traffic fatalities. Prediction models estimated that mortality rates due to COVID-19 infection, in the absence of mitigation, would reach 3,000 deaths per day in England & Wales by the end of May 2020, and over 460,000 deaths in total by August,^12^ a much higher burden of death than observed in this study for both COVID-19 and non-COVID-19 related mortality.

## Findings

Around a quarter of excess deaths during the study period did not have a diagnosis of COVID-19 infection recorded on the death certificate. This may be partly attributable to missed diagnoses, for example where testing did not occur, there was a false negative result or infection did not produce noticeable symptoms.

The risk of missed diagnosis is likely to have reduced over time, as capacity for testing has increased, the tests have become more accurate and clinicians were encouraged to record suspected COVID-19 infection on death certificates.^13^ This may even have resulted in bias in the opposite direction, with deaths misattributed to COVID-19 infection later in the outbreak, which may partly explain the negative values for non-COVID-19 excess deaths in the week ending 8 May. Nevertheless, our results suggest that focusing the health service and wider society on managing a single disease has had unintended consequences. Creating capacity within the health system to manage anticipated demand from patients infected with COVID-19 necessitated the cessation of ‘non-essential’ activity - in reality, non-urgent activity. In addition to reduced supply, there was a simultaneous reduction in demand from patients seeking to protect health services and avoid exposure. As a result, attendances in A&E departments in England in April 2020 were 57% lower than in 2019 and emergency hospital admissions were 39% lower,^14^ while access to health services for people with pre-existing conditions was 20% lower during the COVID-19 peak period.^15^ Non-attendance may have serious immediate consequences, with a reduction in high acuity attendances requiring urgent admission such as acute coronary syndromes, strokes and heart failure. The focus on protecting the NHS also exposed those dependent on the social care sector,^16^ particularly high-risk residents living in close proximity in care homes. Both COVID-19 and non-COVID-19 deaths rose rapidly in these settings,^17^ accounting for over half of all excess deaths in England and Wales.

The observed differences in impacts of COVID-19 across the regions of England and Wales have a range of potential causes that will need to be explored in further detail. COVID-19 spreads through close human contact, and high rates of infection quickly developed in the major conurbations, particularly London, Birmingham (West Midlands), Liverpool and Manchester (North West), and this is reflected in regional excess mortality rates. Following exposure, there are several key risk factors for adverse outcomes, including age, ethnicity, multi-morbidity (particularly diabetes, cardiovascular disease, and conditions and treatments suppressing the immune system), smoking and obesity. However, with respect to age, the regions with the lowest – rather than the highest – proportion of the population over the age of 65 (London, the West Midlands and the North West) had the highest rates of excess deaths, although older populations within the regions were most affected. London is an outlier in terms of age, with only 12.1% of the population over age 65 and 1.7% over age 85 compared to 18.5% and 2.5% respectively for England and Wales, but it was disproportionately affected by the pandemic in its early stages. With respect to ethnicity, Wales and the North East have the smallest BAME populations and London by far the highest.^18^ Other key risk factors, including obesity, smoking and multimorbidity are strongly associated with deprivation,^19 20^ the highest concentrations of which are found in the North East, North West,^21^ and the south of Wales.^22^ Regional rates of excess deaths were not clearly associated with levels of deprivation, although within regions more deprived communities have been disproportionately affected,^2^ reflecting higher prevalence of risk factors, ability to comply with social distancing requirements and exposure to air pollution.^23^

To date, trends in non-COVID-19 related excess deaths during the pandemic have followed the same general pattern as all cause excess deaths, with some English regions and Wales appearing to be relatively unaffected. This may reflect factors such as differences in: levels of testing, thresholds for diagnosis, sociodemographic structures, levels of rurality, service engagement by older patients, and capacities within local health and social care systems. As the pandemic develops, the balance between COVID-19 and non-COVID-19 related deaths is likely to change, and different patterns may develop across the regions. These patterns will need to be monitored as the longer-term impacts of the pandemic and the public health response emerge.

## Conclusion

Short term trends in excess deaths during the COVID-19 pandemic have not followed typical regional patterns for premature mortality in England and Wales, particularly for deaths not directly attributable to the virus. Some regions where mortality rates tend to be low, for example the South East and East, have had relatively high rates of non-COVID-19 related deaths. Predicting longer-term impacts requires caution, but given the sudden and comprehensive re-prioritisation of services, it is likely that a second wave of non-COVID-19 related need is currently building in the community. When this breaks it could overwhelm parts of the NHS, and communities with high baseline levels or morbidity are likely to be hit harder than others and this should be recognised in planning post-pandemic health and social care services.^24^ The national response will need to address local variations in viral activity, in health and social care provision, and in pandemic preparedness, including management of ‘routine’ care during a national emergency. It will also need to address long-standing structural disadvantages that expose parts of the country to avoidable harm from both chronic and acute infectious disease.

## Data Availability

All data used in the study is publicly available from the UK Office for National Statistics

https://www.ons.gov.uk/peoplepopulationandcommunity/birthsdeathsandmarriages/deaths/datasets/weeklyprovisionalfiguresondeathsregisteredinenglandandwales

